# Cohort profile: A multicenter evaluation of clinical decision rules applied to emergency department triage of patients presenting with acute respiratory infection or infectious diarrhea

**DOI:** 10.1101/2023.10.12.23296964

**Authors:** Simon Berthelot, Maurice Boissinot, Michel G. Bergeron, Marie-Louise Vachon, Sylvie Trottier, Ann Huletsky, Rodica Gilca, Jason Robert Guertin, Cécile Tremblay, Yves Longtin, Marc Afilalo, Éric Mercier, Ève Dubé, David Simonyan, Mahukpe Narcisse Ulrich Singbo, Ariane Bluteau

## Abstract

**Purpose:** Emergency department (ED) patients suffering from acute respiratory infection or infectious diarrhea often present with self-limiting conditions. The study objective was to evaluate the performance of triage clinical decision rules consisting of a rapid molecular test and a self-administered patient questionnaire to identify ED patients who can self-treat at home without consulting an emergency physician. This article describes the profile of the cohorts recruited.

**Participants:** Participants were prospectively recruited in 4 EDs in Québec City and Montréal, Canada, from February 2022 through March 2023. Participants were aged ≥18 years, had an acute respiratory infection and/or acute infectious diarrhea, and had received a Canadian Triage and Acuity Scale score between 3 (urgent) and 5 (non-urgent). Participants were asked to complete a self-administered risk stratification questionnaire after triage and to follow usual ED care afterward. Nasopharyngeal and/or rectal swabs were collected and frozen for subsequent testing on a rapid molecular testing device. Data were obtained during the recruitment visit, during a follow-up phone call 7 days later and from medical records. The primary outcome to be predicted by the clinical decision rules was an aggregation of hospitalization, return visit and mortality at 7 days.

**Findings to date:** We recruited 1,391 participants, 62.3% of whom were women, 80.7% were aged under 60, 78.2% had no comorbidities, 76.5% presented with an acute respiratory infection, 17.8% with an acute infectious diarrhea and 5.7% with both. Hospitalization and return visits incidence proportions at 7 days were respectively 10.8% and 13.1% for respiratory infections and 14.1% and 16.5% for infectious diarrhea. No death was recorded.

**Future plans:** The data gathered from these cohorts will enable us to test, refine, derive, and validate clinical decision rules used to help ED triage nurses offer the most suitable care to patients presenting with acute respiratory infections or infectious diarrhea.

**Strengths and limitations:** Our study has both strengths and limitations. Among the strengths:

1. The cohorts were recruited from 4 different EDs and reached the target sample size for acute respiratory infections and acute infectious diarrhea.
2. The potential economic impact of the clinical decision rules will be assessed from the perspective of both the health system and the patient.

The main limitations are the following.

1. Cohorts were recruited by convenience sampling and may not be representative of the entire ED population.
2. The patient self-administered questionnaires used in this study were derived from systematic reviews and rapid prototyping, but not according to the methodological standards recommended for the derivation of clinical decision rules. However, the study dataset was built to enable rules to be refined and if necessary, new rules to be derived and internally validated.
3. We recorded a 12.9% loss of participants at the 7-day follow-up phone call. However, the primary outcome measures (return visits, admissions and deaths) will be obtained from provincial administrative databases. These reliable data will enable us to overcome this limitation for future projects to refine and validate robust triage clinical decision rules.

## Introduction

### Acute respiratory infections and acute infectious diarrhea: a burden on emergency departments

According to the Canadian National Ambulatory Care Reporting System, acute respiratory infections such as influenza-like illness and upper respiratory tract infection represent one of the top three causes of emergency department (ED) visits, accounting for 227,935 visits in Canada each year. Similarly, an estimated 19.5 million episodes of acute gastrointestinal illness occur in Canada each year(1), of which 8.8% are seen in an ED or a primary care practice(1). Acute respiratory infections, whether due to influenza-like viruses, SARS-CoV-2, or other respiratory viruses, are usually self-limiting conditions that do not require evaluation or management by a physician, which is true also for infectious diarrhea(2-5).

If patients seeking treatment in an ED for benign viral infections could be identified as low-risk at triage using diagnostic and prognostic tools, they could return home without assessment by a physician or undergoing non-value-adding medical investigation. This approach could significantly reduce the clinical and financial burden of these infections on overcrowded EDs(6-8), while improving the patient experience. However, this would increase the number of tasks for already overworked triage nurses. Attempts to improve ED triage processes without increasing triage nurse workload or direct contact time between triage nurses and patients are numerous(9-11). Patient participation in ED triage using a self-administered evaluation tool has been proposed as a possible solution(12), but data are still too sparse to conclude that such tools are reliable and would improve or facilitate the practice of nursing in the ED patients

A few clinical decision rules or risk complication assessment tools have been developed and in some cases validated(13-17) for predicting the need for medical care or hospital admission of patients with acute respiratory infection or acute infectious diarrhea. To our knowledge, none has been used to identify in ED triage those who could be returned home without medical evaluation, nor has inclusion of a near-patient rapid molecular diagnostic test in decision algorithms or involving the patient in the decision process been studied.

## Objectives

The primary objective of this cohort study is to evaluate the accuracy of two clinical triage decision rules, one for acute respiratory infection and one for acute infectious diarrhea, each consisting of a rapid molecular test and a self-administered patient questionnaire, to identify ED patients who can self-treat at home without consulting a physician. Secondary objectives are: 1) to refine, derive and/or validate diagnostic and risk stratification tools used by care providers to identify low-risk patients at triage; and 2) to describe the clinical and financial burden that acute respiratory infections and acute infectious diarrhea represent in EDs.

## Methods

### Study design and setting

Two cohorts of participants were recruited, one with acute respiratory infection and the other with acute infectious diarrhea. Recruitment by convenience sampling took place prospectively from February 7, 2022 to March 31, 2023 in the EDs of 4 teaching hospitals in Québec (Canada), namely the CHUL and the Hôpital de l’Enfant-Jésus in Québec City, the CHUM and the Jewish General Hospital in Montréal. Together, these EDs account for over 280,000 visits per year and provide general and specialized acute care to the population for a wide range of health conditions.

This study was approved by the CHU de Québec-Université Laval ethics board (MP-20-2022-6152) and is registered in the ClinicalTrials.gov database (NCT05322694).

### Clinical triage decision rules

Figure 1 shows the hypothetical care pathways associated with the application of the rules after formal ED triage. As recommended by the methodological standards for the derivation and validation of clinical decision rules(18), the care pathways of the participants in this study were not altered. Swabs and questionnaires were collected, but retained for later analysis to blind the assessment of clinical decision rule accuracy. The rules are composed of the following tools:

1. BioFire’s syndromic panels to distinguish between a potentially self-limiting viral infection and a bacterial or parasitic infection at higher risk of complications. These panels have demonstrated high sensitivity and specificity(19-22). The respiratory panel detects 18 viruses and 4 bacteria, whereas the enteric panel detects 13 bacteria, 4 parasites and 5 viruses.
2. The web-based app “inFLUence”, a patient self-administered risk-stratification tool developed and refined through a systematic review, user-centered design study and rapid prototyping(23, 24). This tool consists of a series of questions organized into a decision tree. A patient is classified as being at low risk of complications if ALL questions assessing disease severity (e.g., shortness of breath at rest) or identifying significant comorbidities (e.g., diabetes) and medication (e.g., chemotherapy) are answered “no”. A single “yes” elevates the risk. The self-administered questionnaire for patients with acute infectious diarrhea (inFLUence-GI) was adapted from the respiratory version, but not validated before this study.

**Figure 1.**
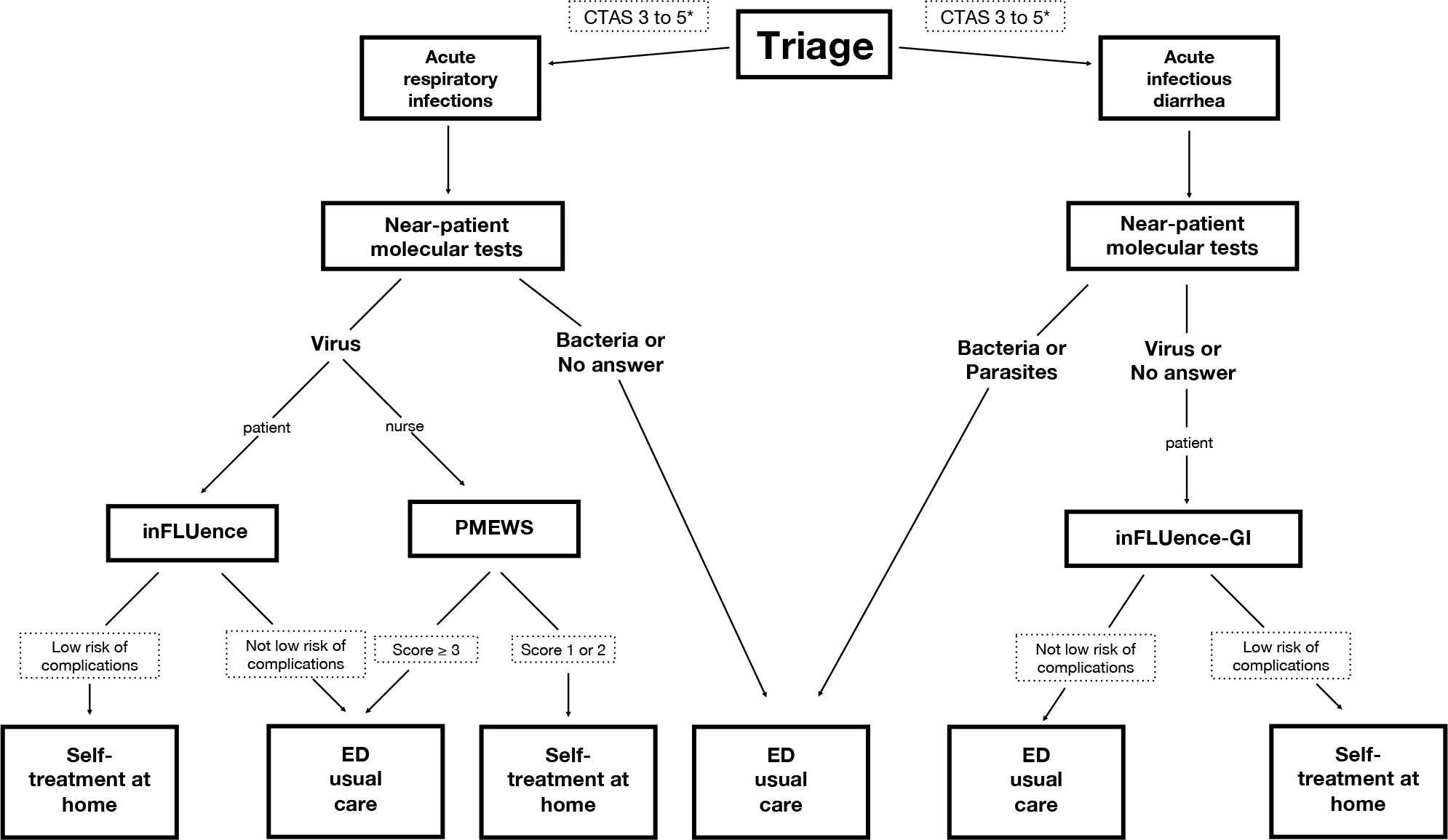
Clinical decision rules and hypothetical care pathways.

To assess the reliability and accuracy of the respiratory questionnaire, an alternative risk-stratification score for acute respiratory infections was also obtained, namely the Pandemic Medical Early Warning Score (PMEWS). The PMEWS(13, 25-27) is designed for use by care providers to assess the risk of complications in patients with influenza or pneumonia and is based on clinical parameters readily mesurable by the triage nurse.

### Selection of participants

Inclusion and exclusion criteria for each cohort are described in Table 1. Patients meeting the criteria for acute respiratory infection and infectious diarrhea were included in both cohorts.

**Table 1.**
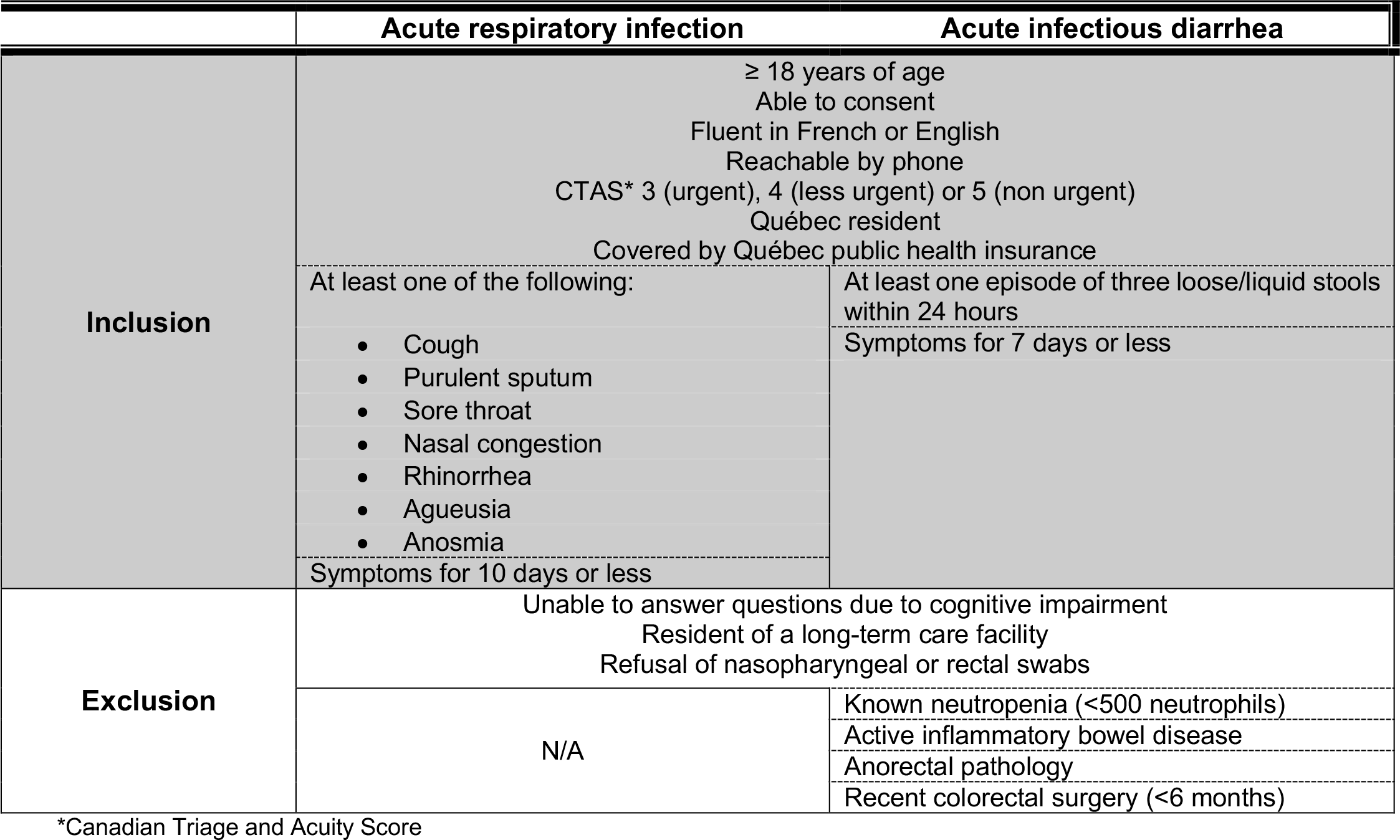
Inclusion and exclusion criteria for acute respiratory infection and acute infectious diarrhea.

### Research procedures

Triaged patients identified as potential participants using the local ED Information System were approached by research staff who confirmed eligibility, obtained consent, collected the demographic and clinical data required for the study, performed the nasopharyngeal or rectal swabbing when applicable, and guided the participants to enable them to complete the risk stratification tool questionnaire themselves via a secure web link on a dedicated research tablet.

Swabs were frozen for subsequent analysis with the BioFire RP2.1 panel or the GI panel. Performed in real time and in proximity to the patient, the test has a turnaround time of about 1 hour. For ED triage purposes, rectal swabbing offers a more convenient and quicker way to obtain an enteric specimen compared to collecting a stool sample but is an off-label use of the GI panel.

Once all study procedures were completed, participants were directed back to the usual ED care pathway that they would have followed normally had they not participated in the study. The molecular test and risk stratification results were not disclosed to the patient, to the care team or to the research staff.

A follow-up was conducted by phone 7 days after the initial visit to determine whether any complications (unplanned return to the ED or a clinic, hospitalization, or death) had occurred, or whether any medication (e.g., antibiotics, antivirals) had been prescribed since the initial visit. The Cost for Patient Questionnaire (CoPaQ)(28) was also administered at this time. The CoPaQ is a validated questionnaire that identifies the expenses paid by the patient (e.g., parking fees) related to the initial visit and the indirect costs of his or her illness (e.g., loss of income due to absence from work).

### Outcome measures

The primary and secondary outcome measures are described in Table 2. These allowed us to note 1) any clinical event (admission, return visit, medical treatment, death) suggesting that evaluation by the emergency physician would have been better advised, and 2) indicators to estimate the operational (ED length of stay) and economic impact (costs) of a new care pathway for patients discharged after triage without seeing the physician.

**Table 2.** Primary and secondary outcome measures.

|  | Acute respiratory infection | Acute infectious diarrhea |
| --- | --- | --- |
| **Primary** | Unplanned return visit to the ED*, unplanned visit at a clinic, hospitalization, or death due to the infection within 7 or 30 days |  |
| **Secondary** | Unplanned return visit to the ED, unplanned visit at a clinic, hospitalization, or death, considered separately at 7 days and at 30 days<br>Admission into intensive care at 7 days and 30 days<br>Length of initial visit at the ED<br>Cost of the illness from the patient's perspective on the 7<sup>th</sup> day<br>Cost of the initial ED visit from the health system perspective<br>Antibiotic prescriptions at 7 days |  |
|  | Medical treatments including i) prescription of antibiotics or antivirals upon discharge from the ED or ii) treatment for acute asthma or chronic obstructive pulmonary disease prescribed during the ED stay or upon discharge from the ED | Medical treatments including i) prescription of antibiotics upon discharge from the ED or ii) intravenous rehydration during the ED stay |
|  | Antiviral prescriptions at 7 days (e.g., oseltamivir) | N/A |

\*Emergency department

### Data sources and measurement

The following demographic data and baseline clinical characteristics were obtained either directly from the participant during the initial visit, by phone during the day 7 follow-up, or from medical records: age, sex and gender, ethnicity, triage score, triage vital signs, initial ED orientation (stretcher, waiting room, redirection to outpatient clinics), comorbidities, regular medication, SARS-CoV-2 and influenza vaccination status, and postal code (deprivation index)(29, 30). The PMEWS was calculated using clinical data collected during the initial visit and was not communicated to the patient or to the care team at that point.

Outcome measures (return visit, hospitalization, death, antiviral or antibiotic prescriptions, ED length of stay) were obtained over the phone on day 7 and by reviewing medical records. Investigations and treatments administered during the ED stay were also recorded in the medical records for later use to estimate the cost of care from the health system perspective. A request for access to the databases kept by the *Institut de la statistique du Québec* has been submitted. These data should become available in early 2024 and will be used to validate the responses obtained during the follow-up call; 2) to identify with more certainty, from medical billing, hospital discharge summaries and death registries, the participants who returned to an ED or went to a clinic, were admitted to hospital, or died within 7 or 30 days of the initial visit; 3) to obtain a deprivation index for each participant; and 4) to refine the economic analysis.

All data were collected on REDCap, a secure web-based platform for creating and managing online databases. To assess the interobserver reliability of medical record extraction, 10% of medical records were extracted independently by two different research assistants and Kappa statistics were calculated.

### Sample size

Assuming a proportion of 10% of patients falling into the combined outcome category of unplanned return visit to the ED or at a clinic, hospitalization, or death at 7 and 30 days, it may be estimated that 47 such patients would be needed to detect a change in the sensitivity of the triage clinical decision rule from 50% (null hypothesis, pre-study) to 70% (alternative hypothesis, post-study) with a sample size of 470 patients, a power of at least 80% and an expected statistical significance level of 0.05. Given the uncertainty associated with changes in the pre-pandemic epidemiology of respiratory viruses, it was determined that at least 1100 participants would allow valid analysis stratified by age and other clinically relevant variables.

## Cohort description

### Characteristics of the participants

Figure 2 shows the number of patients approached and included, as well as the reasons for exclusion. Overall, 31.2% of those approached were recruited for the study. Table 3 describes their demographic and clinical characteristics. Of the 1,391 participants, 62.3% were female, 80.7% were aged under 60, 76.5% presented with an acute respiratory infection, 17.8% with an acute infectious diarrhea and 5.7% with both, 78.2% had no comorbidities, and 78.9% had received at least 2 doses of the COVID-19 vaccine.

**Table 3.**
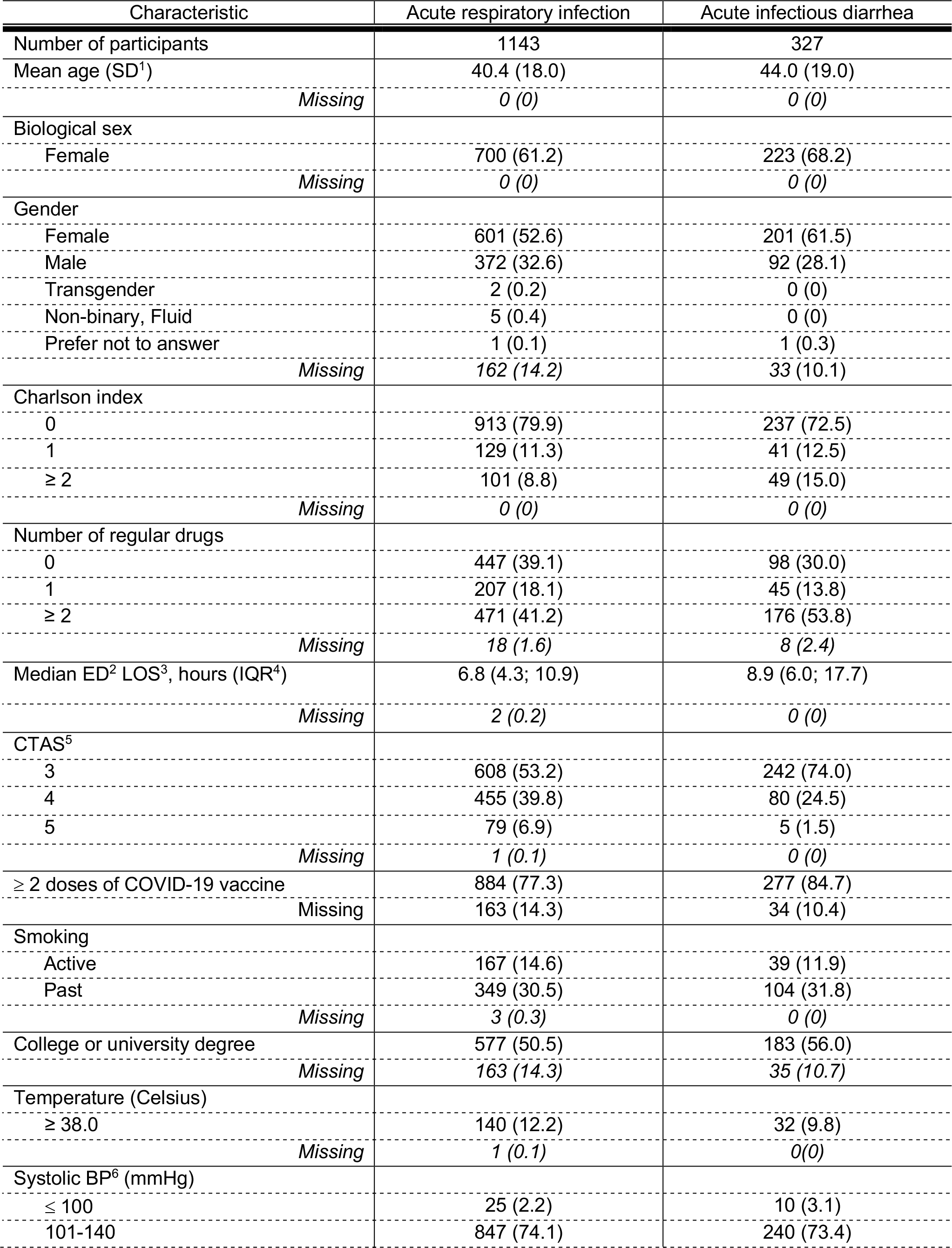
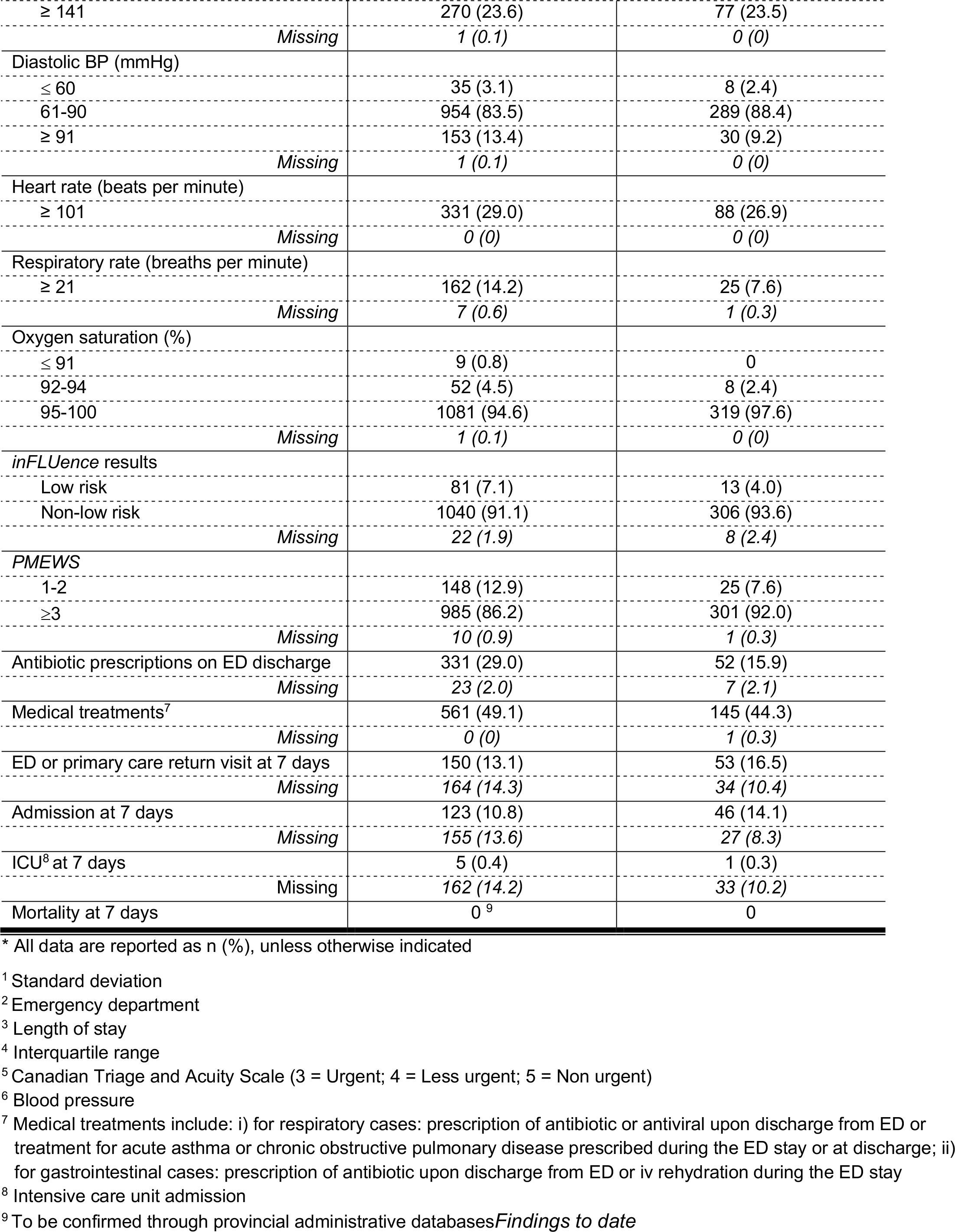
Characteristics^*^ of the participants.

**Figure 2.**
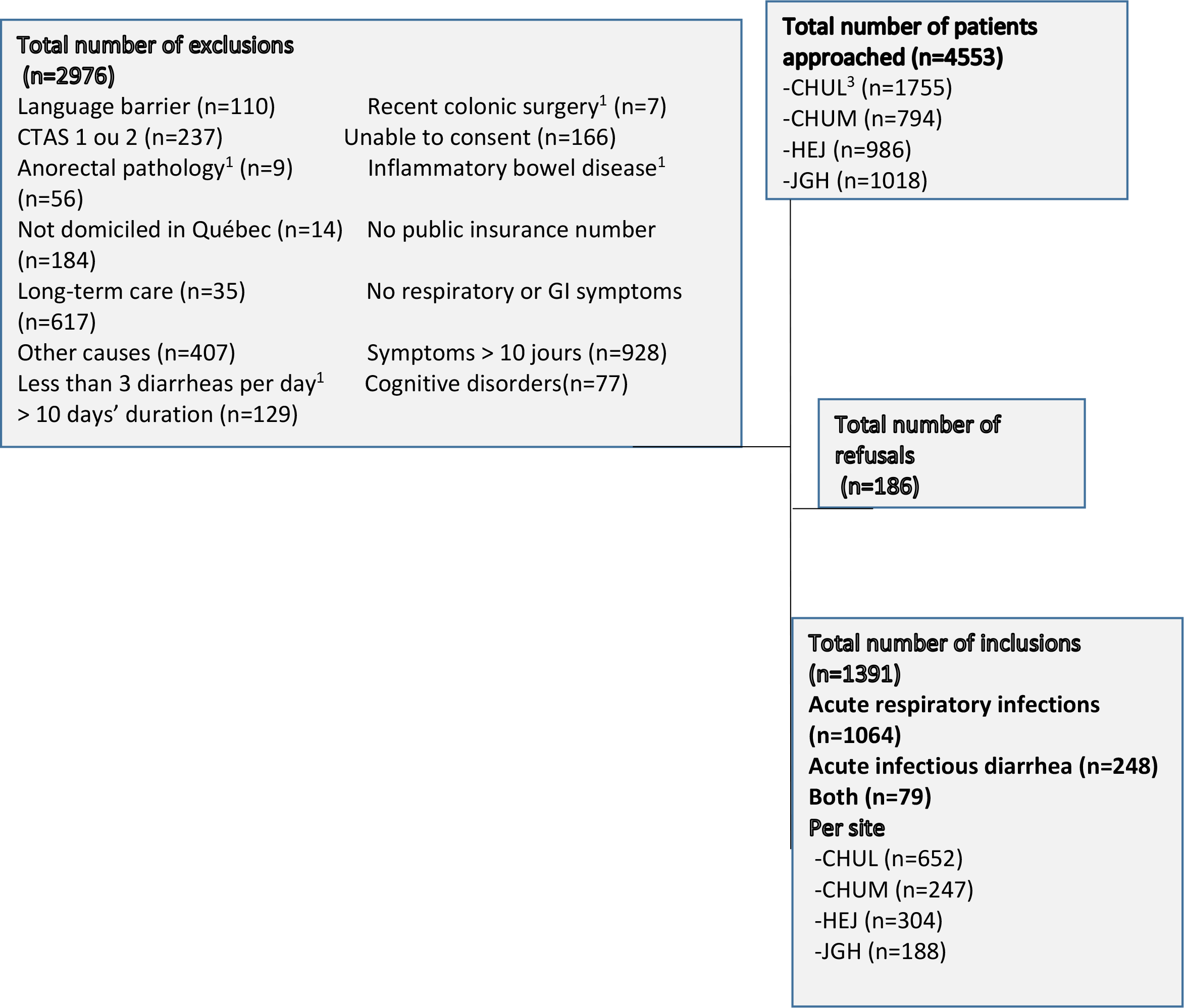
Flow diagram of participants approached, excluded and recruited. ^1^Exclusion criteria only for cohort with acute infectious diarrhea ^2^Canadian Triage and Acuity Scale ^3^Participating hospitals: CHUL, Centre hospitalier de l’Université de Montréal (CHUM), Hôpital de l’Enfant-Jésus (HEJ), Jewish General Hospital (JGH)

Interobserver agreement between two independent research assistants on variables extracted from the medical records of 10% of all participants showed a median kappa statistic of 0.82 (IQR: 0.65; 0.98).

Results for the outcome measures are reported in Table 3. The incidence proportions of hospitalization and return visits at 7 days were respectively 10.8% and 13.1% for acute respiratory infections and 14.1% and 16.5% for acute infectious diarrhea. Moreover, 49.1% of participants suffering from an acute respiratory infection received medical treatment either during their ED stay or were discharged with a prescription, whereas this proportion was 44.3% for those with acute infectious diarrhea.

The inFLUence questionnaire classified 7.1% of participants as being at low risk of developing complications in the case of acute respiratory infection, and 4% in the case of acute infectious diarrhea. Similarly, the PMEWS classified 12.9% of acute respiratory infections and 7.6% of acute infectious diarrhea as low risk.

BioFire panel results for both infections are presented in Tables 4 and 5. Overall, 629 participants (55.0%) in the acute respiratory infection cohort and 157 (48.0%) in the acute infectious diarrhea cohort had at least one identified pathogen, with respectively 4.6% and 12.5% having 2 to 6 pathogens. All respiratory infectious agents identified were viruses, SARS-CoV-2 being the most prevalent. The most frequent enteric pathogen was Norovirus.

**Table 4.**
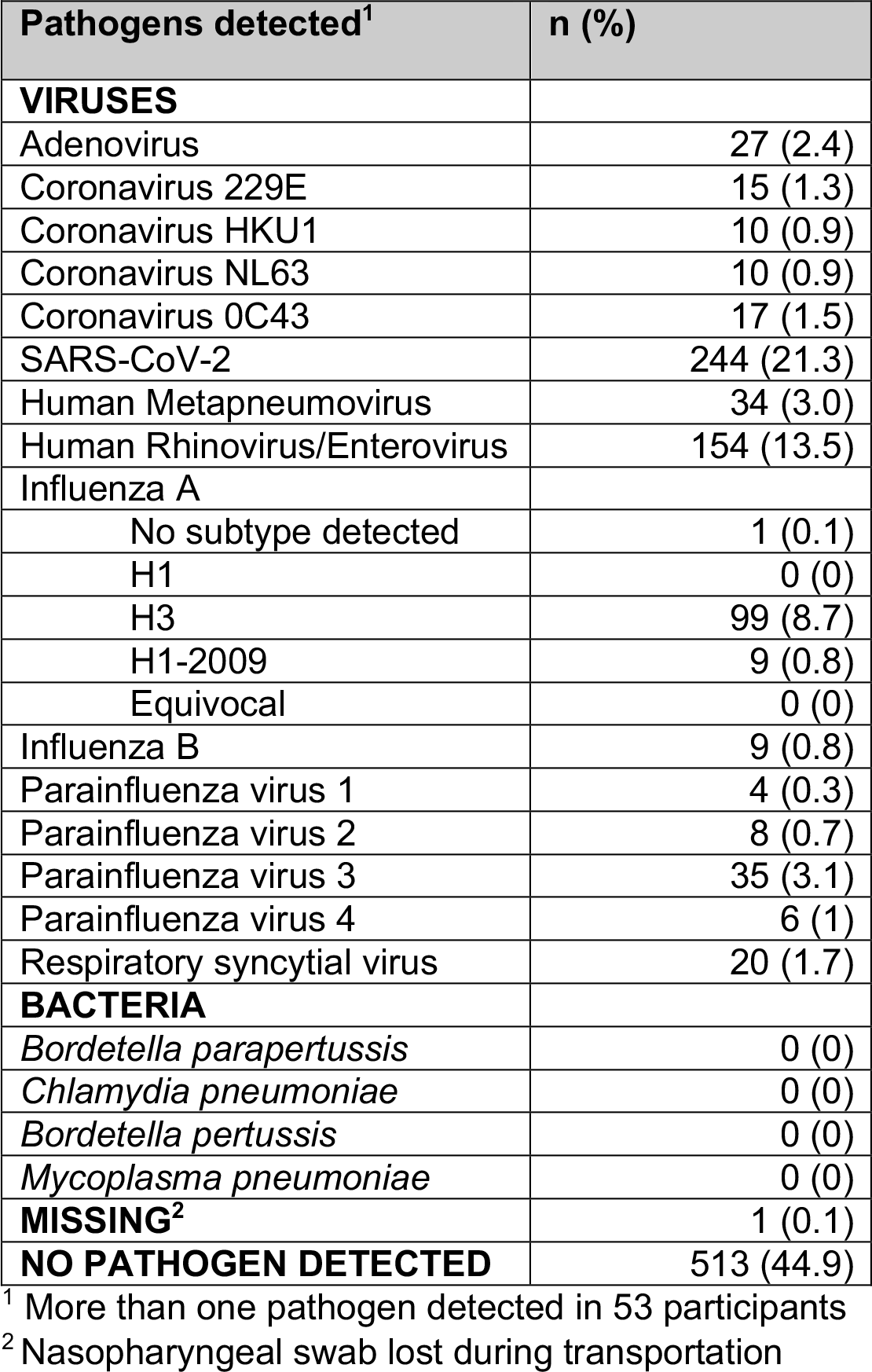
Pathogens detected by the BioFire Respiratory 2.1 Panel (n = 1143 participants)

| Pathogens detected<sup>1</sup> | n (%) |
| --- | --- |
| **VIRUSES** |  |
| Adenovirus | 27 (2.4) |
| Coronavirus 229E | 15 (1.3) |
| Coronavirus HKU1 | 10 (0.9) |
| Coronavirus NL63 | 10 (0.9) |
| Coronavirus OC43 | 17 (1.5) |
| SARS-CoV-2 | 244 (21.3) |
| Human Metapneumovirus | 34 (3.0) |
| Human Rhinovirus/Enterovirus | 154 (13.5) |
| Influenza A |  |
| No subtype detected | 1 (0.1) |
| H1 | 0 (0) |
| H3 | 99 (8.7) |
| H1-2009 | 9 (0.8) |
| Equivocal | 0 (0) |
| Influenza B | 9 (0.8) |
| Parainfluenza virus 1 | 4 (0.3) |
| Parainfluenza virus 2 | 8 (0.7) |
| Parainfluenza virus 3 | 35 (3.1) |
| Parainfluenza virus 4 | 6 (1) |
| Respiratory syncytial virus | 20 (1.7) |
| **BACTERIA** |  |
| *Bordetella parapertussis* | 0 (0) |
| *Chlamydia pneumoniae* | 0 (0) |
| *Bordetella pertussis* | 0 (0) |
| *Mycoplasma pneumoniae* | 0 (0) |
| **MISSING<sup>2</sup>** | 1 (0.1) |
| **NO PATHOGEN DETECTED** | 513 (44.9) |

<sup>1</sup> More than one pathogen detected in 53 participants
<sup>2</sup> Nasopharyngeal swab lost during transportation

**Table 5.** Pathogens detected by the BioFire GI Panel (n = 327 participants)

| Pathogens detected<sup>1</sup> | n (%) |
| --- | --- |
| **VIRUSES** |  |
| Adenovirus F40/41 | 8 (2.4) |
| Astrovirus | 9 (2.8) |
| Norovirus GI/GII | 56 (17.1) |
| Rotavirus A | 7 (2.1) |
| Sapovirus (I, II, IV, and V) | 5 (1.5) |
| **BACTERIA** |  |
| *Campylobacter* (*C. jejuni* / *C. coli* / *C. upsaliensis*) | 16 (4.9) |
| *Clostridioides* (*Clostridium*) *difficile* (toxin A/B) | 26 (8.0) |
| *Plesiomonas shigelloides* | 1 (0.3) |
| *Salmonella* | 5 (1.5) |
| *Yersinia enterocolitica* | 0 (0) |
| *Vibrio* (*V. parahaemolyticus* / *V. vulnificus* / *V. cholerae*) | 0 (0) |
| *Vibrio cholerae* | 0 (0) |
| Enteraggregative *E. coli* (EAEC) | 17 (5.2) |
| Enteropathogenic *E. coli* (EPEC) | 29 (8.9) |
| Enterotoxigenic *E. coli* (ETEC) lt/st | 6 (1.8) |
| Shiga-like toxin-producing *E. coli* (STEC) stx1/stx2 | 5 (1.5) |
| *E. coli* O157 | 0 (0) |
| *Shigella*/Enteroinvasive *E. coli* (EIEC) | 6 (1.8) |
| **PARASITES** |  |
| *Cryptosporidium* | 6 (1.8) |
| *Cyclospora cayetanensis* | 2 (0.6) |
| *Entamoeba histolytica* | 0 (0) |
| *Giardia lamblia* | 2 (0.6) |
| **MISSING** | 0 (0) |
| **NO PATHOGEN DETECTED** | 170 (52.0) |

<sup>1</sup> More than one pathogen detected in 41 participants

## Future plans

The data collected from these cohorts with acute respiratory infection and/or acute infectious diarrhea will enable us to complete the following projects:

1. Evaluate the accuracy and performance (sensitivity, specificity, likelihood ratios, ROC) of near-patient molecular testing and patient self-administered risk-stratification tools as clinical decision rules for triage nurses in EDs (the primary goal of the present cohort study).
2. Derive and internally validate through secondary analyses two new clinical decision rules based on assessment by triage nurses to help identify patients that need to see an emergency physician.
3. Estimate the potential economic benefit, to the patient and to the health system, of implementing the new clinical decision rules derived and validated in the previous steps (patients at low risk of complications sent home to self-treat without medical assessment). The cost to patients will be assessed using the CoPaQ(31), a validated questionnaire that estimates costs absorbed by patients and caregivers for a given illness. The cost to the health system will be assessed by applying a time-driven activity-based costing methodology that has been adapted for use in the ED by our team and described in detail elsewhere(32).
4. Using the same costing methods, compare the cost of care, from the patient and health system perspectives, for patients with COVID-19 versus other respiratory infections, and for patients with enteric bacterial infections versus viral etiology.
5. Evaluate and compare molecular testing systems to routine testing by the care team for respiratory infections and diarrhea, especially the accuracy of enteric panels with off-label use of rectal swabs.
6. Assess the PMEWS as a predictor of hospital admission and mortality in a population for which it has not been studied, namely patients with acute infectious diarrhea.
7. Assess sex, gender, race, and socioeconomic status as sources of discrepancies in molecular test results (e.g., SARS-CoV-2), disease severity (e.g., PMEWS), patient-borne costs (CoPaQ) and outcome (e.g., hospitalization).
8. Determine the proportion of respiratory infections caused by Enterovirus D-28 (specific genotype unidentified by commercial multiplex PCR panel) and compare its severity to that of other respiratory infections using PCR, genomics, and statistics.
9. Using data collected from interviews of patients and care providers, identify barriers to and facilitators of implementing clinical decision rules to discharge patients who, based on triage, do not require medical care. The results of this qualitative study will be published in parallel.

## Strengths and limitations

### Our study has both strengths and limitations. Among the strengths

3) The cohorts were recruited from 4 different EDs and reached the target sample size for acute respiratory infections and acute infectious diarrhea.
4) Access to provincial databases will provide us with reliable data on key outcome measures (return visits, hospitalization and death).
5) The rich research database including near-patient molecular test results will enable us to refine our clinical triage decision rules and explore many other secondary research questions related to the overall scope of the project. This will be performed using standard analytical/statistical approaches as well as artificial intelligence.
6) The potential economic impact of the clinical decision rules will be assessed from the perspective of both the health system and the patient.

### The main limitations are the following

4) Cohorts were recruited by convenience sampling and may not be representative of the entire ED population.
5) The patient self-administered questionnaires used in this study were derived from systematic reviews and rapid prototyping, but not according to the methodological standards recommended for the derivation of clinical decision rules. However, the study dataset was built to enable rules to be refined and if necessary, new rules to be derived and internally validated.
6) Near-patient molecular tests were not performed in real time. Swabs were frozen for later testing due to the lack of accessibility of the testing device. Although a pilot study has demonstrated the feasibility of a bedside testing approach in the ED, this multicenter study cannot confirm whether the use of molecular testing at the patient’s bedside at triage is feasible in a real clinical context.
7) We recorded a 12.9% loss of participants at the 7-day follow-up phone call. However, the primary outcome measures (return visits, admissions and deaths) will be obtained from provincial administrative databases. These reliable data will enable us to overcome this limitation for future projects to refine and validate robust triage clinical decision rules.

## Collaboration

De-nominalized demographic and clinical data will be made available to qualified researchers upon request. To protect participant confidentiality, this will not include sensitive data such as postal codes. We encourage collaborations that align with the goals of our research and may enhance our understanding of the findings of this cohort study. Potential collaborators are invited to discuss research proposals and opportunities for joint analyses or other scientific cooperation. Access to data will be subject to certain restrictions and data-sharing protocols to comply with ethical guidelines. Researchers seeking access to the data will be required to sign a data-sharing agreement outlining the terms and conditions of data use and confidentiality.

## Conclusion

We have described the cohorts of a multicenter study conducted in 4 teaching hospitals in Québec (Canada) from February 2022 through March 2023 and aimed at improving the care pathway for patients with acute respiratory infection or acute infectious diarrhea who may be directed to self-care at home after triage in the ED. We have amassed a large dataset on these patient populations and tested new ED triage tools, including diagnostic near-patient molecular tests and patient self-administered risk stratification questionnaires. The cohorts recruited for the two targeted conditions will enable us to test, refine, derive and/or validate clinical decision rules to help ED triage nurses and care providers offer these categories of patient the care most appropriate for their condition

## Patient and public involvement statement

Patients or the public were not involved in the design, or conduct, or reporting, or dissemination plans of our research

## Declaration of funding

Funding for this study was received from the *Fonds d’accélération des collaborations en santé* of the *Ministère de l’Économie et de l’Innovation du Québec*.

## Data availability

The data sets generated during and/or analyzed during this study are available from the corresponding author on reasonable request.

## Notes

### Competing Interest Statement

The research team received funding from Meridian Bioscience to develop PCR assays. The BioFire assays used in this study were duly purchased from Biomerieux. However, none of these companies had influence over study design or on any of the data published in this manuscript.

### Clinical Trial

NCT05322694

### Funding Statement

This study was funded by the Ministere de l'economie et de l'innovation du Quebec

### Author Declarations

Ethics committee/IRB of Centre de recherche du CHU de Quebec-Universite Laval gave ethical approval for this work

